# The maintenance of complex visual scenes in working memory may require activation of working memory manipulation circuits in the dlPFC

**DOI:** 10.1101/2023.11.11.23298415

**Authors:** Frederick Nitchie, Abigail Casalvera, Marta Teferi, Milan Patel, Kevin Lynch, Walid Makhoul, Yvette Sheline, Nicholas L Balderston

## Abstract

Past research has shown that the bilateral dorsolateral prefrontal cortices (dlPFC) are implicated in both emotional processing as well as cognitive processing,^1,2,3^ in addition to working memory^4, 5^. Exactly how these disparate processes interact with one another within the dlPFC is less understood. To explore this, researchers designed an experiment that looked at working memory performance during fMRI under both emotional and non-emotional task conditions. Participants were asked to complete three tasks (letters, neutral images, emotional images) of the Sternberg Sorting Task under one of two trial conditions (sort or maintain). Regions of interest consisted of the left and right dlPFC as defined by brain masks based on NeuroSynth^6^. Results showed a significant main effect of the ‘sort’ condition on reaction speed for all three trial types, as well as a main effect of task type (letters) on accuracy. In addition, a significant interaction was found between trial type (sort) and task type (letters), but not for either of the picture tasks. These results reveal a discrepancy between BOLD signal and behavioral data, with no significant difference in BOLD activity during image trials being displayed, despite longer response times for every condition. While these results show that the dlPFC is clearly implicated in non-emotional cognitive processing, more research is needed to explain the lack of BOLD activation seen here for similar emotionally valanced tasks, possibly indicating involvement of other brain networks.

## Introduction

Working memory (WM) is defined as the ability to mentally store, maintain, and manipulate information during a relatively short period of time^1,2^. Previous studies have found the dorsolateral prefrontal cortices (dlPFC) to play significant roles in working memory functioning^3–6^. This appears especially true in mental health research, where people diagnosed with mental disorders, such as general anxiety disorder and major depressive disorder, show hyperactivity of the dlPFC, which is linked to decreased performance on working memory tasks. The prevailing theory behind such hyperactivation is that those afflicted with mental disorders require greater activation to achieve similar levels of behavioral performance as healthy individuals^7,8^. While the influence of the dlPFC on WM is evident, what is less clear is how the type of content impacts dlPFC involvement.

Previous research suggests that the left and right dlPFC may play discrepant roles in maintaining cognitive and affective information in a person’s working memory^9^. Specifically, the left dlPFC appears primarily involved in processing verbal information, while the right dlPFC is more involved with non-verbal information^10^. This has been demonstrated in verbal n-back tasks while observing BOLD activity in the associated regions, specifically when participants are asked to manipulate spatial information within their working memory^11^. It has also been observed in both behavioral and BOLD imaging studies that the complexity of a performed WM task will show increases in cognitive demand, evidenced by longer response times as well as increases in brain activity. For example, in the Sternberg memory task, which has participants either maintain information as presented or manipulate the information within WM before answering, accuracy is reduced and reaction time is greater in the manipulate condition^12^. In addition to the type of information being processed, there is also evidence that the complexity of processing tasks can also lead to increases in dlPFC BOLD activity^13,14^.

In addition to these findings, bilateral dlPFC has also been implicated in emotional regulation. Lesion studies have shown that patients with damage to their dlPFC are less capable of downregulating fear response to conditioned stimuli than non-lesioned patients^15^. Stimulation to dlPFC with transcranial direct current stimulation (tdcs) is also shown to increase a subject’s ability to regulate their physiological response to an emotionally arousing stimuli as compared to baseline^16^. Indeed, we have proposed that emotion regulation may stem from working memory processes in the dlPFC, with the right dlPFC primarily responding to affective content. Therefore, the purpose of the current study was to determine the extent to which the left and right dlPFC differentially respond to verbal, spatial, and affective content during the Sternberg WM paradigm. Based on previous research, we hypothesized increases in right dlPFC BOLD compared to left for affective content.

## Materials and Methods

### Participants

This cohort consisted of 17 participants recruited from within the University of Pennsylvania and the surrounding community (N = 17). Of those 17, seven identified as male, nine identified as female, and one identified as non-binary. Ages ranged from 18 – 57 (xL = 34.06, σ = 11.26). Participants were included if they were: (i) between 18 and 60 years of age; (ii) demonstrated normal cognition; (iii) able to read and understand English; (iv) right-handed; (v) were found by investigators to have no history of meeting DSM criteria for any diagnosis; (vi) and were able to provide informed consent. Participants were excluded if they met any of the following conditions: (i) were currently pregnant; (ii) were unable to tolerate, or else were medically or surgically contraindicative to MR scanning procedures; (iii) had a history of stroke, epilepsy, or brain scarring; (iv) showed signs of cognitive impairment; (v) were determined by investigators to have recently used any psychoactive medication(s). All participants signed an informed consent form, and the protocol was approved by the Institutional Review Board for human subject research at the University of Pennsylvania. The authors assert that all procedures contributing to this work comply with the ethical standards of the relevant national and institutional committees on human experimentation and with the Helsinki Declaration of 1975, as revised in 2008.

## Procedure

Participants who met basic eligibility criteria were consented to the study and escorted to the scanner. Subjects were prepped to go into the scanner, de-metaled, given ear plugs, situated comfortably, and given the response device and emergency squeeze ball. Once in the scanner, structural scans were collected followed by the 3 task runs.

## Materials

### Sternberg Working Memory

The Sternberg task employed in this study required subjects to either sort or maintain items from 3 categories: letters, neutral images, and emotional images from the Open Affective Standardized Image Set (OASIS) database^17^. Neutral images included images items, scenes, and people. Emotional images included mutilated bodies, injuries, fires, vehicular accidents, and other highly arousing negative images. Valence and arousal ratings from the OASIS norms were used to select negatively valanced vs neutral images. Each trial started with an instruction keyword to indicate the trial type. Next, subjects viewed a series of 5 items (letters, neutral images, emotional images), presented sequentially, and instructed to retain them in working memory for a brief retention interval. On half of the trials (maintain trials) they were instructed to remember the items in the order presented. On the other half of the trials (sort trials) they were instructed to rearrange the items. Subjects rearranged letters in alphabetical order and images in reverse order. After the retention interval, subjects were presented with an item and a number and instructed to indicate whether or not the position of the item in the series (original or sorted depending on trial type) matched the number. Half of the trials were matches, half were mismatches. The duration of the letter series (1.5 – 2.5 s), retention interval (3.5 – 5.5 s), and ITI (3 – 8 s) were jittered across trials. The duration of the instructions (1 s) and response prompt (3 s) were fixed. Subjects completed 3 runs of the task with 24 trials per run and 12 per condition.

### Scans

Scans were acquired on a 3 Tesla Siemens Prisma scanner with a 64-channel head coil (Erlangen, Germany). Scans included a T1, T2, and 3 fMRI task runs. The T1-weighted scan was an MPRAGE (TRL=L2200Lms; TEL=L4.67Lms; flip angleL=L8°) with 160, 1 mm axial slices (matrixL=L256L ×L256; field of view (FOV)L=L240LmmL×L240Lmm). The T2-weighted scan (TRL=L3200Lms; TEL=L563Lms; flip angleL=Lvariable) included 160, 1 mm sagittal slices (matrixL=L256LmmL×L256Lmm; FOVL=L240LmmL×L240Lmm). For each task, we acquired 615 whole-brain BOLD images (TRL=L800 ms; TEL=L37Lms; flip angleL=L52°; Multi-band acceleration factor = 8) comprised of 72, 2 mm axial slices (matrixL=L104L×L104; FOVL=L208LmmL×L2008Lmm) aligned to the AC-PC line.

### fMRI Pre-processing

Task data were processed using the afn_proc.py script distributed with the AFNI software package ^18^. We used the following preprocessing blocks: `tshift`, `align`, `volreg`, `blur`, `mask`, `scale`, `regress`. These steps accomplished the following standard preprocessing steps: 1) slice-time correction, 2) EPI/T1 alignment using an Local Pearson Correlation cost function, 3) volume registration (reference volume = the image with the fewest outliers), 4) blurring with a 2 mm Gaussian kernel, 5) masking with the intersection of the EPI brain mask and the skull-stripped T1, 6) scaling so the mean of the run was 100, 7) censoring (scrubbing) of images with greater than 0.5 mm displacement or greater than 15% of voxels registered as outliers, 7) timeseries regression using the 6 primary motion vectors and their derivatives, and a set of polynomial regressors to model the baseline. Task regressors corresponding to the encoding, retention, and response period of the WM trials were modelled using variable duration blocks to account for the jittering of trial events.

## Statistical Analyses

### Performance

Accuracy (percent correct) and reaction time was calculated for each individual and each condition. Trials with no response trials were counted as missing for reaction time scores. These scores were analyzed using a 3 (task: letters vs. neutral images, vs. emotional images) by 2 (trial: sort vs. maintain) repeated measures ANOVA. Interaction effects were characterized by paired sample t-tests where appropriate. Partial eta-squared and Cohen’s d were calculated for ANOVA effects and t-tests, respectively.

### dlPFC BOLD

BOLD activity during the retention interval was extracted from regions of interest encompassing the left and right dlPFC. These regions were defined using NeuroSynth (Yarkoni et al., 2011), which was used to generate a bilateral mask of the dlPFC. We completed a search with the term ‘dlPFC’, saved the resulting association test map and extracted the primary clusters corresponding to the left and right dlPFC. We then averaged the BOLD values for the retention interval across voxels within each dlPFC region. Finally, we performed a 2 (hemisphere: left vs. right) x 3 (task: letters vs. neutral images, vs. emotional images) by 2 (trial: sort vs. maintain) repeated measures ANOVA on BOLD values extracted from the dlPFC.

## Results

### Sternberg performance

#### Accuracy

To determine the effect of stimulus type on working memory performance, we performed a 3 (task: letters vs. neutral images, vs. emotional images) by 2 (trial: sort vs. maintain) repeated measures ANOVA on accuracy and reaction time scores. For accuracy, we observed a significant main effect of task (f(2,30) = 8.91; p = 0.001; eta-squared = 0.37), but no main effect of trial (f(1,15) = 0.63; p = 0.44; eta-squared = 0.04) and no task by trial interaction (f(2,30) = 0.24; p = 0.788; eta-squared = 0.02).

To characterize the main effect, we conducted *post hoc* t-tests comparing the accuracy across the tasks. Subjects were significantly more accurate for letter vs. both neutral (t(15) = 3.51; p = 0.003; d = 0.88) and emotional (t(15) = 3.80; p = 0.002; d = 0.95) images, which did not differ from each other in terms of accuracy (t(15) = 0.42; p = 0.684; d = 0.11).

#### Response time

For response time, we observed a main effect for trial (f(1,15) = 6.28; p = 0.024; eta-squared = 0.29), but no main effect of task (f(2,30) = 0.41; p = 0.669; eta-squared = 0.03) and no task by trial interaction (f(2,30) = 0.09; p = 0.91; eta-squared = 0.01), suggesting that subjects were slower for sort compared to maintain trials, independent of task type.

#### dlPFC BOLD

To understand whether the tasks evoked similar patterns of dlPFC-related BOLD, we performed a 2 (hemisphere: left vs. right) x 3 (task: letters vs. neutral images, vs. emotional images) by 2 (trial: sort vs. maintain) repeated measures ANOVA on BOLD values extracted from the dlPFC. We found a significant main effect for hemisphere (f(1,16) = 4.83; p = 0.043; eta-squared = 0.23) with greater BOLD responses in the left dlPFC compared to the right. We also found a significant trial by task interaction (f(2,32) = 3.76; p = 0.034; eta-squared = 0.19), but no other main effects or interactions (all ps > 0.05)

To characterize this interaction, we performed a series of *post hoc* sort vs. maintain t-tests for the letters, neutral images, and emotional images separately. As in previous work, we found significantly greater BOLD activity for sort trials vs. maintain trials in the letter task (t(16) = 2.91; p = 0.01; d = 0.71). In contrast, we found no difference for either the neutral (t(16) = 0.53; p = 0.607; d = 0.13) or the emotional (t(16) = 0.49; p = 0.63; d = 0.12) images. In both cases, a *post hoc* power calculation suggests that we would need at least 368 subjects to reliably detect an effect.

## Discussion

In this experiment, healthy participants were given multiple versions of the Sternberg working memory task while undergoing fMRI. The content differed across versions, with three content types presented: letters, neutral images, or emotionally arousing images. On each trial, subjects either maintained the items in WM or sorted the items in WM (alphabetically for trials, or reverse order for the image trials). For our primary analysis, we extracted BOLD activity in the left and right dlPFC. Counter to our initial hypothesis, we did not find a laterality effect for either the neutral or emotional images. However, for both image types we found an increase in bilateral dlPFC activity during the maintenance condition compared to the letter condition. These results suggest that the maintenance of visual information in WM relies heavily on dlPFC activity.

### BOLD activity

The primary finding from the BOLD data stem from a task x trial interaction. For letters, our results replicate the common effect that working memory manipulation drives dlPFC activity. However, when comparing working memory maintenance to manipulation for both the neutral and emotional images, we see no difference between in BOLD activity in the dlPFC. Indeed, it seems that dlPFC BOLD seems to be elevated during both the maintain and sort trials. These results suggest that even the maintenance of complex visual scenes requires the active manipulation of information in working memory. Accordingly, we argue that subjects were engaged in some form of elaborative verbal working memory strategy aimed at rapidly encoding the gist of each complex scene. Although speculative, this verbal encoding hypothesis suggests that adding a concurrent verbal task should interfere this task, which is testable. Another interesting implication of the current findings is that image maintenance is a simple effective tool to drive dlPFC activity that could be used to manipulate context during neuromodulation interventions ^19^.

Critically, we must also discuss our findings with respect to the original hypotheses. Initially, we had hypothesized a task x hemisphere interaction, rather than a task by trial interaction. Specifically, we expected to find greater left dlPFC activity for verbal content and greater right dlPFC activity for non-verbal content. This hypothesis was based on previous work using the n-back task that generated content-specific BOLD laterality effects in the dlPFC ^11^. Similar effects have been shown using neuromodulation, suggesting that stimulating right dlPFC interferes with the verbal n-back, while stimulating the left dlPFC interferes with spatial n-back^20^. The primary difference between these studies and the current work is the complexity of the visuospatial information used. The previous studies required subjects to maintain the spatial location of letters in working memory, rather the complex visuospatial information in rich visual scene.

In addition to the interaction, we found an overall main effect of hemisphere, with greater BOLD signal in the left dlPFC across all versions of the task. Previous studies have shown differential hemispheric activation when participants are tasked with manipulating verbal, as opposed to non-verbal, information during fMRI; with greater BOLD activity being shown in the left dlPFC as compared to the right, which was an effect we saw replicated in this cohort^10,21^.

These results are consistent with the previous argument that subjects were relying on verbal rather than spatial strategies to maintain the images in working memory, which also could explain our lack of laterality effects. Perhaps future studies can use dual task designs to probe the maintenance of visual information in WM while reducing reliance on verbal strategies.

Our finding of increased left dlPFC activity during letter sorting replicates previous work showing an increase in BOLD activity when cognitive load increases ^22,23^. In contrast, we did not replicate previous work showing right dlPFC activity increases for associated with the increase in visual working memory load during the sort condition for the image trials ^24,25^. One interesting finding of note is that seeming discrepancy between dlPFC BOLD and reaction time (discussed in detail below), where, despite showing no change in either left or right dlPFC activity during sort trials with complex images, participants still displayed significant increases in their response times on these trials, suggesting less efficient processing on these trials ^26^. Future research should investigate how task difficulty and executive processes are mediated across the brain ^27^.

### Response time

As expected, we found a main effect of trial type on response time with ‘sort’ trials taking longer on average to respond to, compared to ‘maintain’ trials, with participant responding faster on maintain trials as compared to sort trials. This finding was significant across all task types, supporting the conclusion that working memory manipulation imposes additional task demands regardless of the content type. Although just a manipulation check, these results support future work using the current version of the task. This finding is also consistent with the attentional control theory, which suggests that as cognitive demand is increased (in this case through categorically increasing or decreasing the complexity of a given trial), more effort is required to complete task requirements, and therefore processing efficiency is decreased^26^.

These results agree with many findings in the literature, demonstrating that as cognitive load increases due to task complexity, participants will take longer to answer trials on a given task. (Altamura, Veltman, etc.)

Consistent with previous literature, as cognitive load increases due to task complexity, participants will take longer to answer trials on a given task^14,28^. Interestingly, we did not observe a significant difference in response time as a function of stimulus type. This seems to run counter to previous work suggesting that tasks involving complex visual stimuli require longer reaction times, and that these reaction times are further increased for negatively valenced stimuli ^29,30^. This inconsistency further supports our hypothesis that subject used verbal strategies to order the pictures in this task.

### Accuracy

We found a main effect of task on accuracy, revealing that participants were more accurate when responding to tasks using letters as compared to either neutral or emotional images. We did not find any significant effect of trial type, with participants showing no difference in performance between maintain and sort trials in terms of accuracy across task types (verbal, neutral image, emotional image). This makes some intuitive sense, as letters are more familiar than the novel images presented to the participants, and one would expect maintaining (or sorting) the more familiar stimuli to be somewhat easier. In addition, an inherent aspect of using letters is that they already have a set order that nearly all participants would be expected to have memorized. This could potentially aid in arranging them correctly on either trial type as compared to the images, which were arbitrarily ordered in their conditions. It is noteworthy that we did not see a difference between the emotional images and the neutral ones, suggesting – consistent with the verbal strategy hypothesis above – that emotional content did not negatively impact performance.

Our findings here showing fewer errors for non-visual vs visual stimuli are consistent with previous experiments focusing on similar behavioral tasks. This finding seems to hold even when load is experimentally manipulated, with a high load non-visual task still resulting in higher accuracy compared to a low-load visual task ^31^. While this finding is consistent with previous literature, we did not see differences in accuracy for negative vs. neutral images, as has been previously shown ^30,32^. One possible explanation for this lack of emotion effect is that the ordering task potentially triggered a verbal retention strategy, thus stripping the images of their negative valence. Finally, we did not observe any difference in accuracy as a function of task demands (i.e. maintain vs. sort) ^12,28^, one likely explanation is that subjects took more time to complete the manipulation trials, prioritizing accuracy over reaction time.

### Strengths and Limitations

There were several strengths to the current study. First, we used a well-controlled and extensively researched working memory paradigm, that has been repeatedly shown to drive dlPFC activity. Second, our primary analysis was based on an *a priori* hypothesis and relied on *a priori* regions of interest, thus limiting the possibility of obtaining false positives in the fMRI data. Third, although not technically a strength, the current work raises interesting and novel testable hypotheses for future studies. Despite these noted strengths, it should be noted that the study included a relatively small sample size of 17 participants in total. Additionally, the results were counter to our hypotheses. Although not technically a limitation, this work should be replicated in a larger independent sample.

## Conclusions

Here we showed the dlPFC increases activity during the manipulation of verbal but not visual information. One interpretation of these results is that we rely on complex manipulation of verbal content in working memory to encode rich visual scenes. Although this conclusion is only inferred from the fMRI data and needs replication, it raises interesting testable hypotheses that are important for understanding emotion regulation and for optimizing non-invasive neuromodulation (described above). Future work should explore the use of working memory manipulation as a tool to modulate the context during non-invasive neuromodulation sessions for mood disorders, potentially leveraging content valence to activate, and thus target, specific emotion regulation circuits.

## Author contributions

CRediT author statement according to: https://www.elsevier.com/authors/policies-and-guidelines/credit-author-statement.

**Conceptualization: YIS, NLB**

**Methodology: KGL, YIS, NLB Software: NLB**

**Formal analysis: KGL, NLB**

**Investigation: FN, AC, MT, MP, WM, NLB**

**Writing - Original Draft: FN, NLB**

**Writing - Review & Editing: FN, AC, MT, MP, WM, KGL, NLB**

**Visualization: FN, KGL, NLB**

**Supervision: YIS, NLB**

**Project administration: YIS, NLB**

**Funding acquisition: YIS, NLB**

## Data Availability

All data produced in the present study are available upon reasonable request to the authors

## Acknowledgments

This study utilized the high-performance computational capabilities of the CUBIC computing cluster at the University of Pennsylvania. (https://www.med.upenn.edu/cbica/cubic.html). The authors would like to thank Maria Prociuk for her expertise and assistance in submitting the paper. We would also like to thank the participants for their time and effort.

## Ethical Standards

The authors assert that all procedures contributing to this work comply with the ethical standards of the relevant national and institutional committees on human experimentation and with the Helsinki Declaration of 1975, as revised in 2008.

---

**Figure 1.**
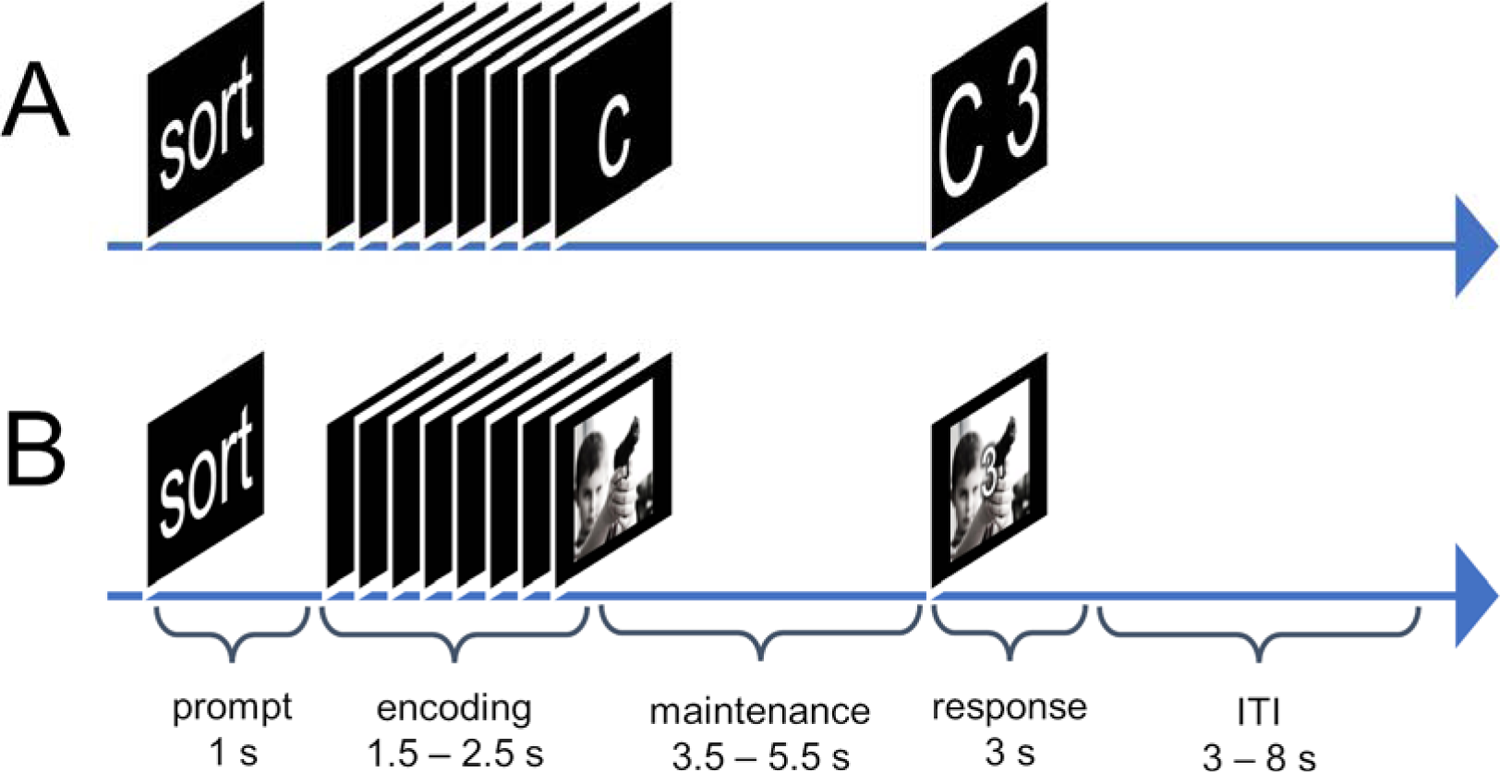
Schematic of task design. A) Example trial from a Sternberg (letter) trial. Subjects were presented with a series of letters and instructed to either (maintain) remember the letters in the order they were presented or (sort) rearrange the letters in alphabetical order. Prior to the trial, subjects were given a prompt that indicated the trial type. After a brief retention interval, subjects were presented with a letter and a number and asked whether the number matched the position of the letter in the original (maintain trials) or rearranged (sort trials) series. **B)** Example from a Sternberg (picture) trial. Subjects were presented with a series of either emotional or neutral images and asked to either sort or maintain the series of images for a brief retention interval. Like the Sternberg Letter task, subjects were presented with a picture and a number and were asked to indicate whether the number matched the position of the picture in the series.

**Figure 2.**
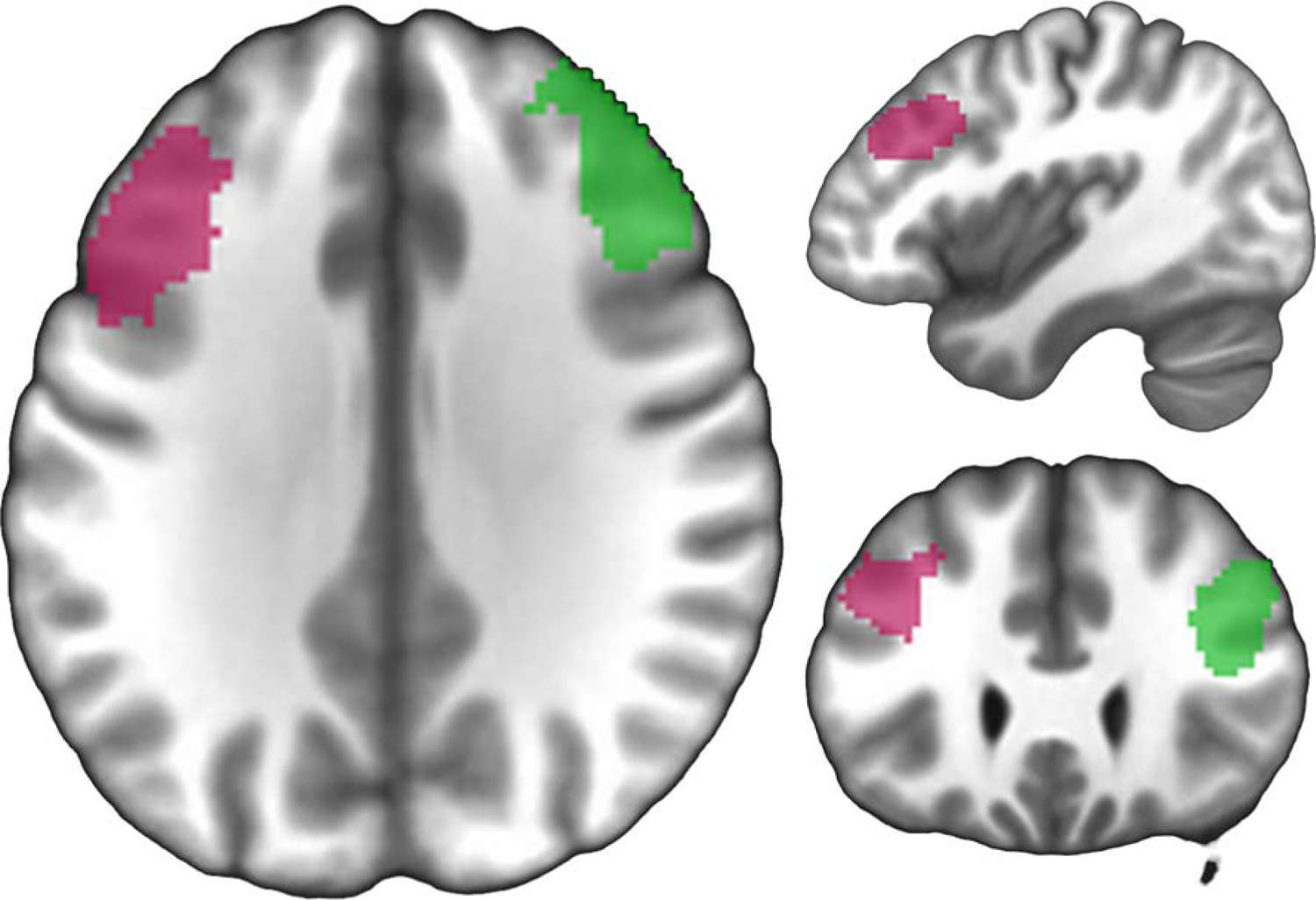
Regions of interest for the left (pink) and right (green) dorsolateral prefrontal cortex (dlPFC). A neurosynth.org (Yarkoni et al., 2011) association test for the term ‘dlPFC’ was used to generate the mask.

**Figure 3.**
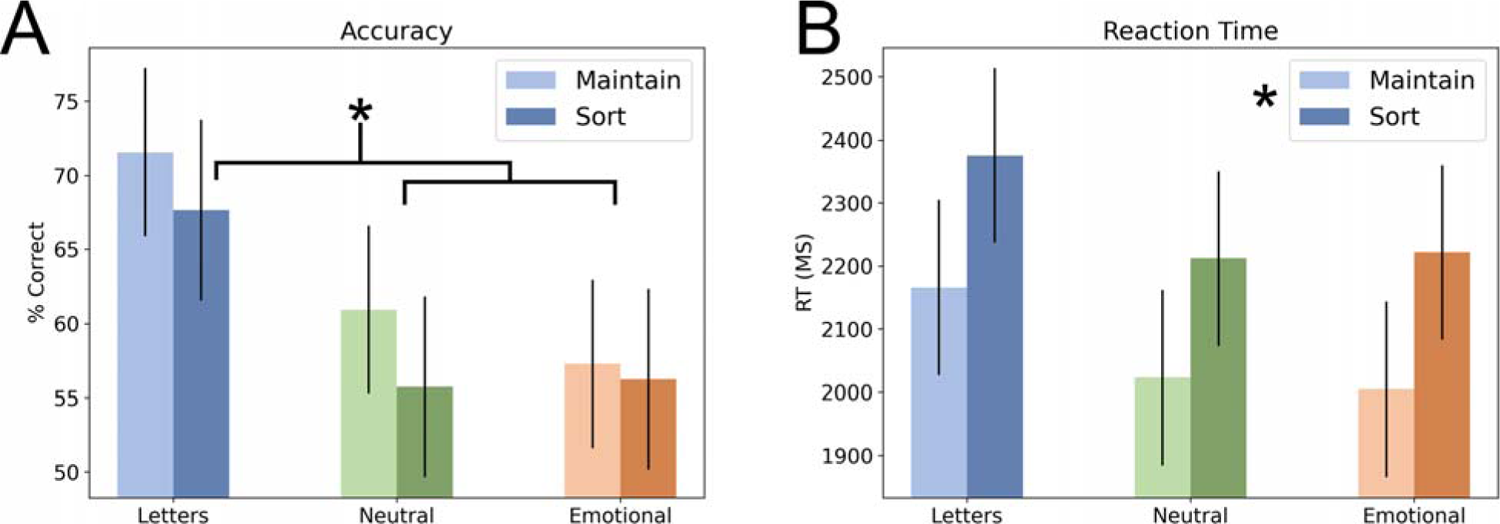
Accuracy and reaction time during the Sternberg WM tasks. A) Percent correct during the task. **B)** Reaction time during the task. Bars represent the mean ± SEM. * = p < 0.05.

**Figure 4.**
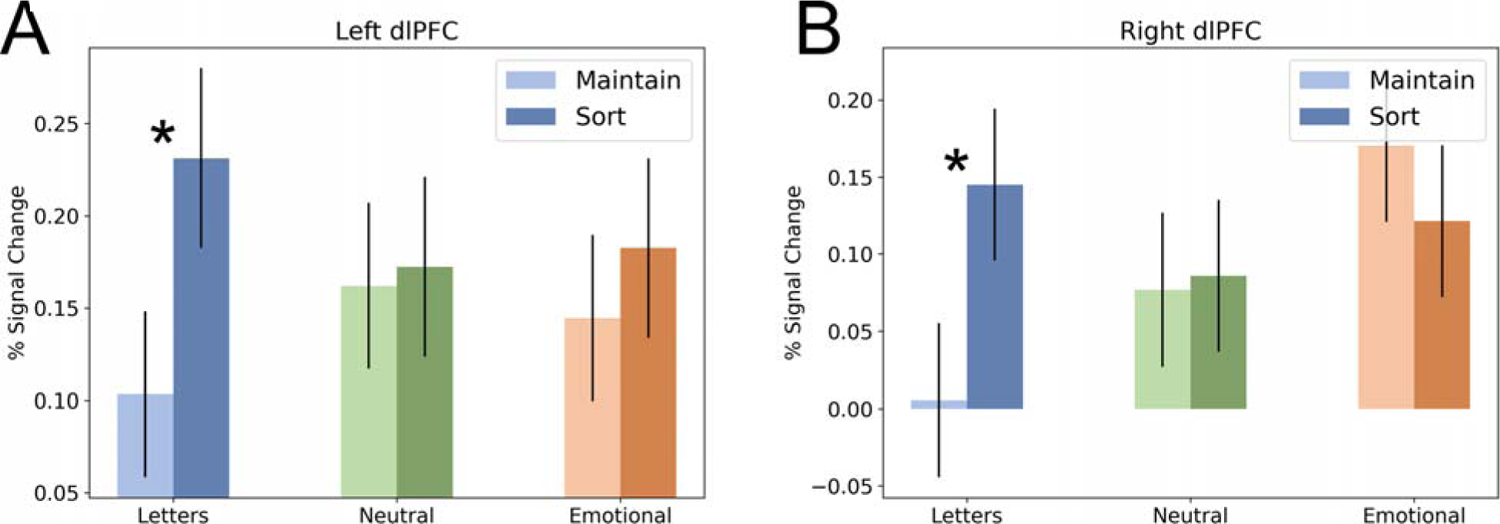
BOLD from the left and right dlPFC regions of interest during the Sternberg WM tasks. A) BOLD from the left dlPFC. B) BOLD from the right dlPFC. Bars represent the mean ± SEM. * = p < 0.05.

